# Epidemic Curve of Contamination in a Hospital That Served as Sentinel of the Spread of the SARS-Cov-2 Epidemic in the City of Rio de Janeiro

**DOI:** 10.1101/2020.10.19.20215079

**Authors:** Marisa Santos, Tereza Cristina Felippe Guimarães, Helena Cramer, Fabiana Mucillo, Izabella Pereira da Silva Bezerra, Raiana Andrade Quintanilha Barbosa, Tais Hanae Kasai Brunswick, Adriana Bastos Carvalho, Glauber Monteiro Dias, Aurora Issa, Adriana Bastos Carvalho, Antonio Carlos Campos de Carvalho

## Abstract

The COVID-19 pandemic had a profound impact on the operation of Brazilian hospital units, even those dedicated to non-infectious diseases. This study aims to describe the Covid-19 epidemic curve from a cardiovascular specialized nosocomial unit. All symptomatic employees were submitted to RT-qPCR. A total of 613 tests were performed on 548 employees between March 23, 2020, and June 4, 2020; with 45.7% positivity from the samples, representing 11.9% of the total employees. The epidemic curve showed a profound drop after first week of May. The data showed a high contamination rate despite widespread availability of personal protective equipment and employees’ training.

## Introduction

The COVID-19 pandemic had a profound impact on the operation of hospital units, even those not dedicated to caring for COVID-19 cases. The need for sick professionals to be absent from work for prolonged periods, the need to create exclusive areas for the treatment of infected patients, the consumption of resources and the fear that employees could act as vectors and contaminate their family and community were the most challenging aspects of operation during the pandemic.

The first case of COVID-19 was described in China on November 17, 2019 (1), although the World Health Organization did not detect the cluster until December 31, 2019 (2). The first case in Brazil was diagnosed on February 25, 2020 (3). The first patient with COVID-19 was admitted to the National Institute of Cardiology on April 8, 2020.

The National Institute of Cardiology is a unit that treats complex cases and serves as a reference center for cardiology in the Ministry of Health. It has 165 beds and approximately 2000 employees and is responsible for approximately 70% of invasive cardiology care in the state of Rio de Janeiro. Heart transplants for adults and children are performed at the institute, and in 2020, it was accredited for lung transplantations.

The present report aims to describe the epidemic curve of contamination for employees of the National Institute of Cardiology during the COVID-19 pandemic.

## Method

### Testing

The testing of employees focused on the early identification of infected individuals; the reduction of cross-transmission to other employees, patients and family members; the reduction of absenteeism; and support for planning the replacement of sick employees.

Employees with any respiratory symptoms, diarrhea, fever, anosmia or myalgia were tested by RT-PCR from the 3rd to the 10th day of disease using nasopharyngeal swabs. The testing included all hospital employees, who can be divided into three categories according to their risk of exposure: those with no contact (administrative personnel), indirect contact (cleaning, information technology, pharmacy, security, general services, building maintenance and clinical engineering personnel) or direct contact with patients (doctors, nurses, physical therapists, nutritionists, and occupational therapists). Asymptomatic patients were not screened. Employees who were positive or had severe symptoms (fever, dyspnea, severe prostration) were removed from work for 14 days.

### RNA extraction

Swab samples were gently homogenized, and 300 µl of transport medium was separated for viral RNA extraction. The rest of the material was stored at -80 °C. Viral RNA was extracted according to the manufacturer’s instructions using the ReliaPrep Viral TNA Mini-Prep kit (Promega, cat. AX4820) or the QIAamp MinElute Virus Spin Kit (Qiagen, cat. 57704).

The samples were subjected to a lysis step consisting of treatment with proteinase K and incubation with lysis buffer at 56 °C for 15 minutes.

Subsequently, a binding buffer was added to the samples, and the mixture was transferred to RNA extraction columns. Three washes with washing buffer were then performed. Finally, the nucleic acids obtained were eluted in 100 µl of nuclease-free water and stored at -80 °C until use.

### Reverse Transcription and Quantitative Polymerase Chain Reaction

Reverse transcription and polymerase chain reaction were performed in a single step using the GoTaq Probe 1-Step RT-qPCR (Promega, cat. A6121) or TaqPath 1-Step RT-qPCR Master Mix kits (Thermo Scientific, cat. A15300). Viral cDNA was detected by a TaqMan assay containing two sense primers and a probe for three independent markers: two detection regions of the viral nucleocapsid (N1 and N2) for human RNAse P (RP). The sequences used were developed by the Centers for Disease Control and Prevention (CDC) in the United States and are sold by Integrated DNA Technologies (cat. 1006713). The assays were performed and analyzed according to the CDC’s instructions. The quantitative PCR equipment used was QuantStudio 5 (Applied Biosystems) and Viia 7 (Applied Biosystems). The tests were considered positive when there was amplification of N1, N2 and RP with a cycle threshold (Ct) below 40. The tests were considered negative when there was amplification of the RP with Ct below 40 and no amplification of N1 and N2 or when there was amplification of both markers with Ct above 40. The tests were considered inconclusive when there was amplification of the RP with Ct below 40 and amplification of only one of the viral markers (N1 or N2) with Ct below 40.

### Statistical analysis

The daily time series of the number of tests performed was analyzed using the unobserved components model (4). The software Stata version 15 (StataCorp, 2017) and the STAMP module of the software OxMetrics version 8 (Jurgen A. Doornik, 2018) were used. The first two weeks corresponded to the period during which the collection process had not yet been fully implemented and testing was irregular; therefore, data from those weeks were excluded from the analysis.

The study was approved by the Research Ethics Committee of the National Institute of Cardiology under number 3234232.,9.0000.5272.

## Results

A total of 613 tests were performed on 548 employees between March 23, 2020, and June 4, 2020; 280 of these tests were positive (45.7% positivity), representing 11.9% of the institute’s total employees. A total of 3.7% of the tests were considered inconclusive and were repeated later.

Table 1 shows the profile of the tested employees.

**Table 1.**
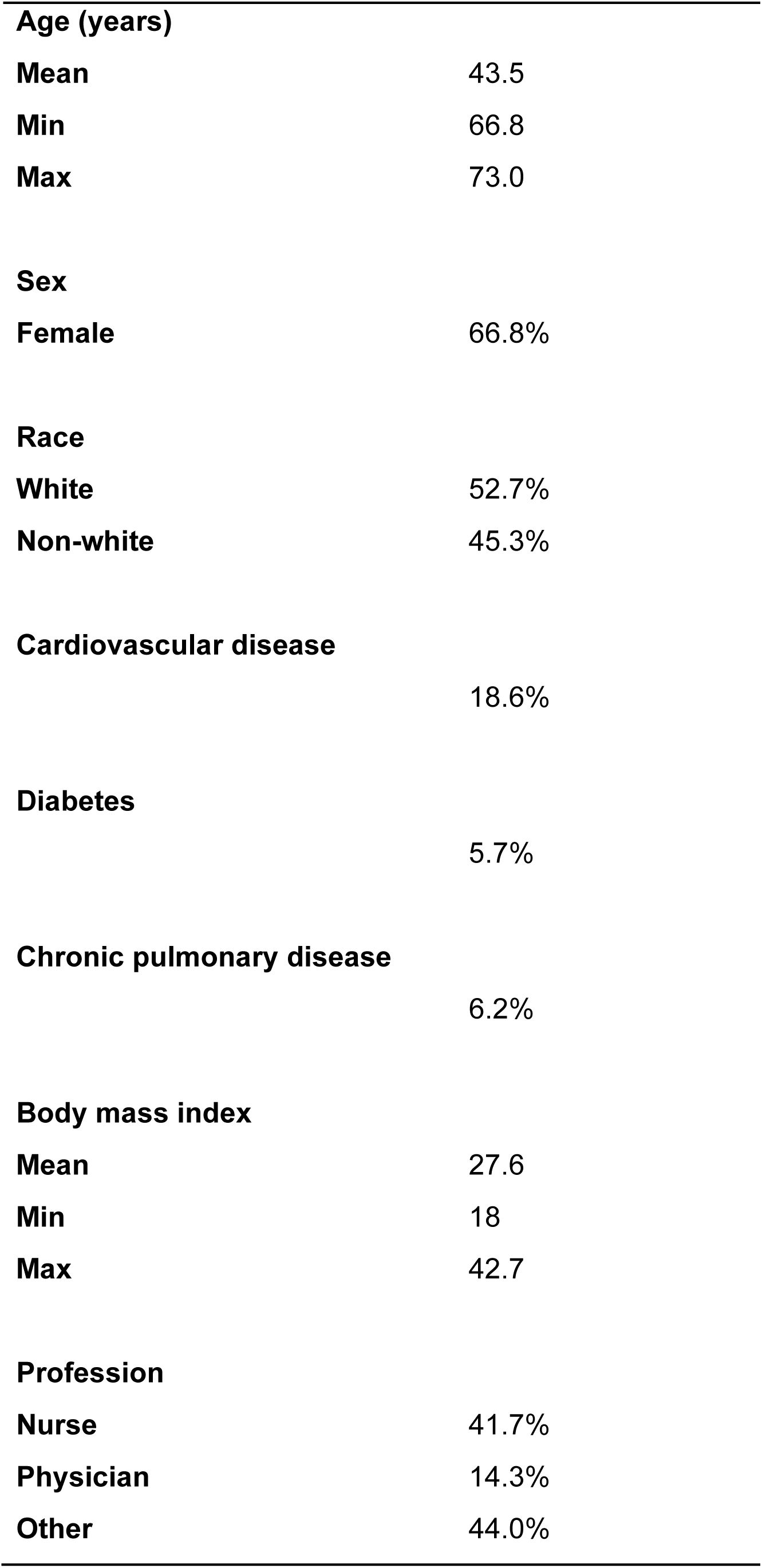
Characteristics of the tested employees

|  |  |
| --- | --- |
| **Age (years)** |  |
| Mean | 43.5 |
| Min | 66.8 |
| Max | 73.0 |
| **Sex** |  |
| Female | 66.8% |
| **Race** |  |
| White | 52.7% |
| Non-white | 45.3% |
| **Cardiovascular disease** |  |
|  | 18.6% |
| **Diabetes** |  |
|  | 5.7% |
| **Chronic pulmonary disease** |  |
|  | 6.2% |
| **Body mass index** |  |
| Mean | 27.6 |
| Min | 18 |
| Max | 42.7 |
| **Profession** |  |
| Nurse | 41.7% |
| Physician | 14.3% |
| Other | 44.0% |

Figure 1 represents the time series with the total number of tested and positive individuals. There was a pronounced drop in the number of tests and the number of positive cases. The positivity rate ranged from 0% (March 23) to 79% (April 28). The curve suggests pronounced growth between April 6 and May 8.

**Figure 1.**
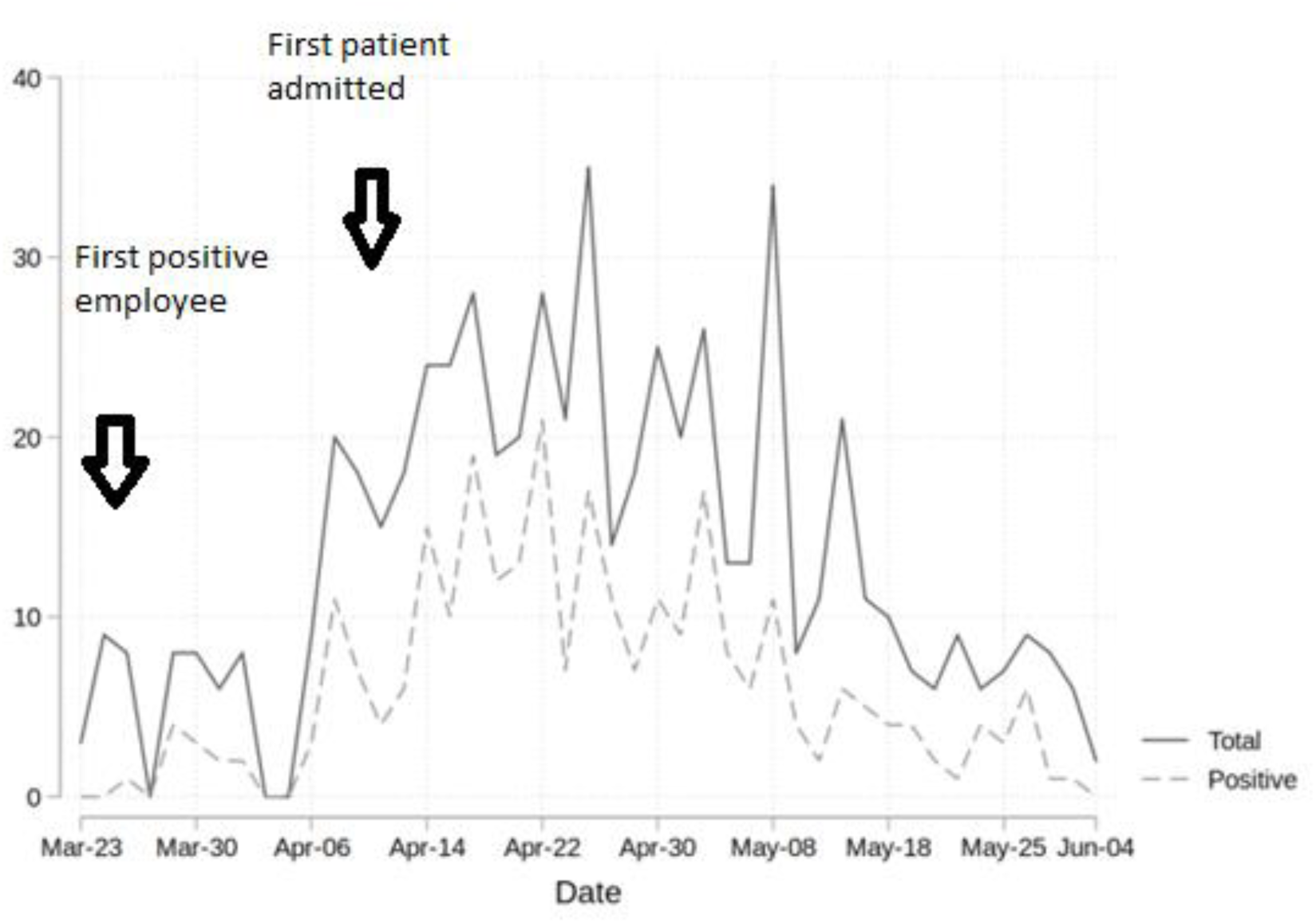
Time curve – total number of tests performed and positive cases per day

The analysis of time series assumes weak stationarity (i.e., the series has a constant mean and variance over time). To stabilize the variance, we applied a logarithmic transformation to the number of tests. Despite the transformation, the first two weeks showed much higher variance than the rest of the series, and it was decided to exclude them from the analysis. Only data from epidemiological weeks 15 to 23 were used, comprising 35 observations (mean of 3.9 observations per week).

Figure 3 represents the logarithm of the series. According to the model, the peak occurred on the 11th day of the time series.

**Figure 2.**
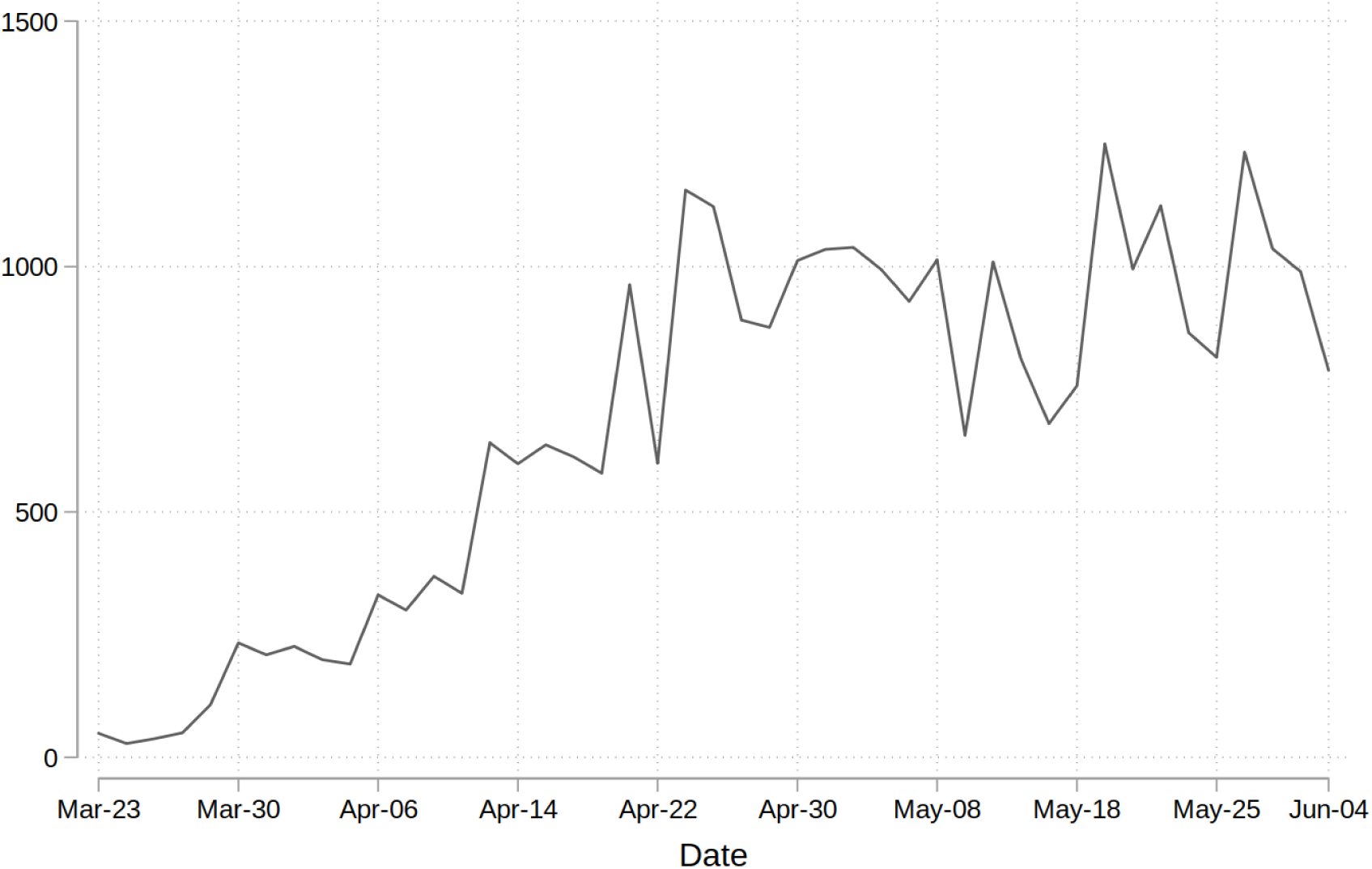
shows the epidemic curve for the municipality of Rio de Janeiro, where the number in infected individuals with an RT-PCR-confirmed diagnosis shows a similar trend but without a pronounced decrease.

**Figure 3.**
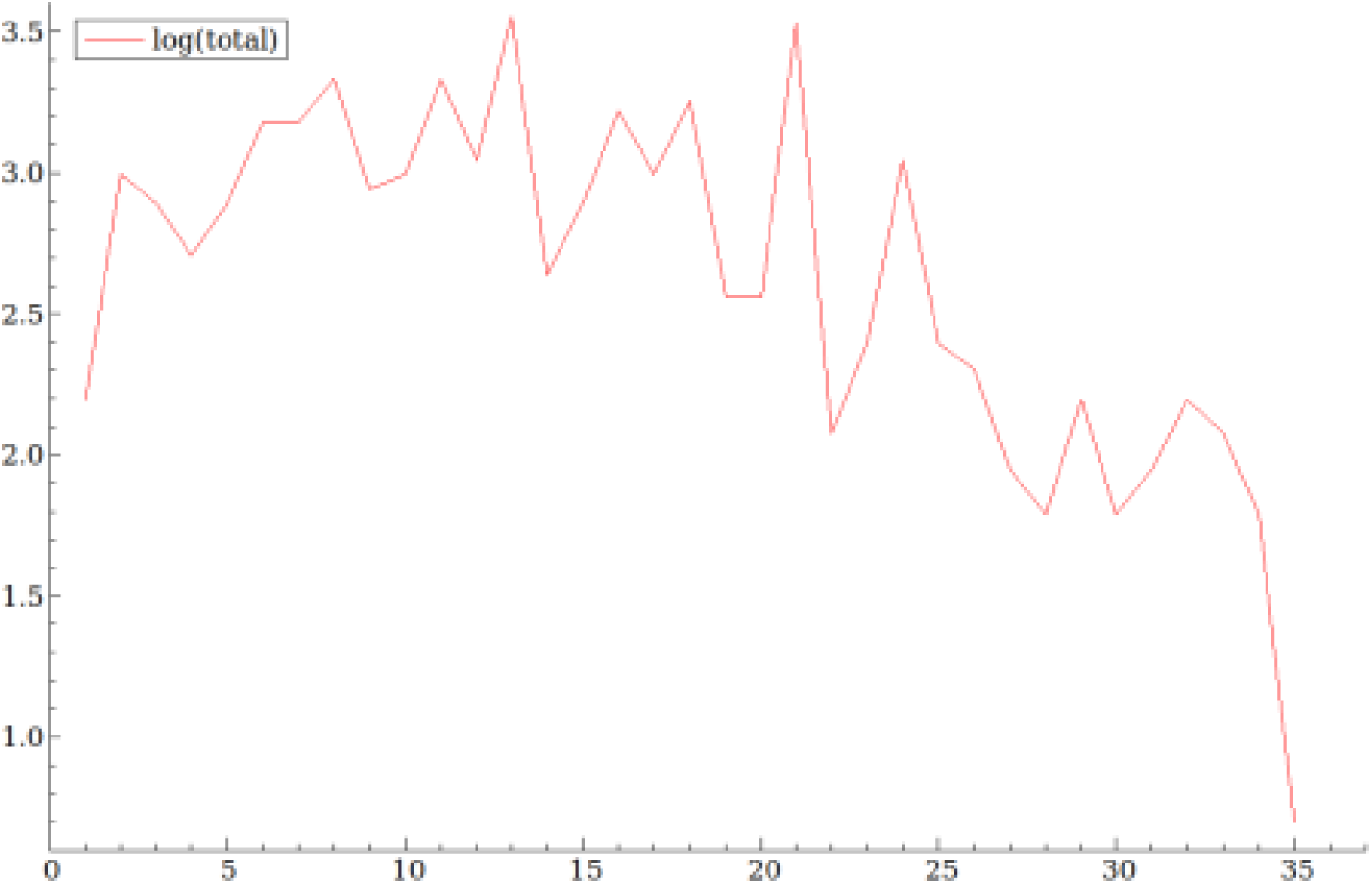
Logarithm of the number of tests/day (x-axis: day of the time series)

Figure 4 shows the four components (trend, seasonality, cycle and error) separately. As seasonal and cyclical components are absent from this series, the local linear trend model was used.

**Figure 4.**
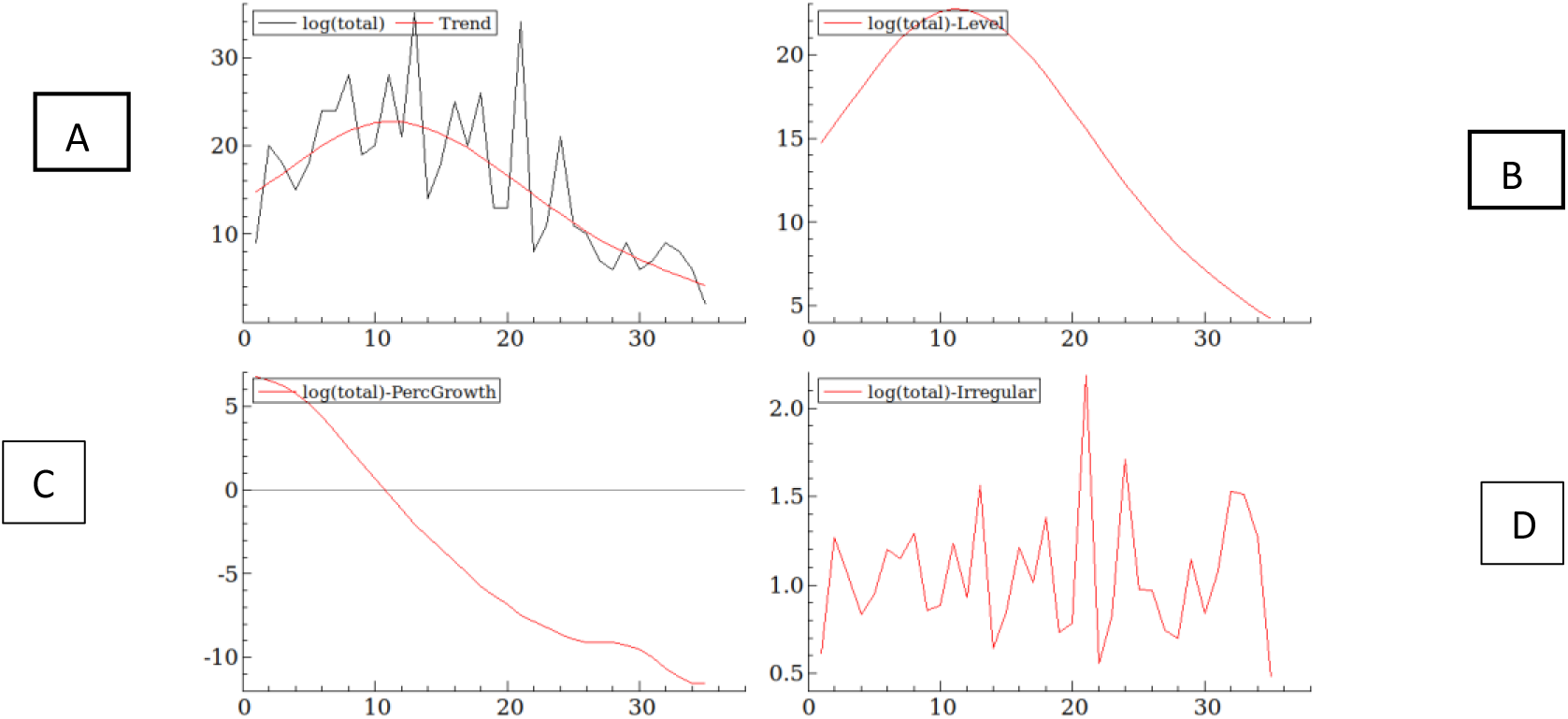
Components of the time series

Graph A shows the level superimposed on the series, graph B shows the level, graph C shows the growth rate, and graph D shows the irregular component or error.

The residuals of the model have an approximately normal distribution (Shapiro-Wilk test p = 0.97) and do not show statistically significant autocorrelation (Ljung-Box test p = 0.29).

## Discussion

An important problem in the analysis of trends and projections of epidemic behavior is the lack of testing. Due to the limited availability of material for testing at the beginning of the epidemic in our country, it was necessary to restrict testing to only symptomatic employees. The hospital represents a microcosm of the city, with employees with various risk profiles, housing areas and socioeconomic statuses, and epidemic behavior in the hospital may mirror that in the city, as indicated by the comparison of the increase in the curves of infected individuals in Figures 1 and 2 (5).

The curve in Figure 1 shows a clear reduction in the number of positive and symptomatic individuals tested after the first week of May at the National Institute of Cardiology, demonstrating a reduction in the infection of employees. In contrast, Figure 2 shows clear stabilization of the number of infected individuals in the municipality of Rio de Janeiro at very high values. The reduction in adherence to social distancing, the migration of the epidemic from smaller geographic regions to those with higher population density and, likely, the temporary re-opening of businesses may explain the changes in epidemic behavior.

Despite the widespread availability of personal protective equipment, the rapid identification of sick employees and the training provided in the hospital, the positivity rate observed until June was considered high compared to that reported in the literature (5–7). Brazil has high rates of COVID-19 contamination and mortality among healthcare professionals, especially nurses (8,9). To date, the cohort has experienced the death of a nursing technician. We speculate that despite the risk of exposure to patients from working in the hospital environment, many employees were contaminated by other employees (asymptomatic, presymptomatic or symptomatic), by family members or when traveling to the hospital by public transport. It is also important to note that professionals work in various public and private hospitals, acting as vectors of transmission between different units. The cases of infection among healthcare professionals at the institute preceded the hospitalization of the first patient, reflecting probable contamination from another unit or from the community. For the vast majority of cases, there are no data to establish where contamination occurred.

An important limitation of this study is the lack of screening of asymptomatic individuals. Some publications estimate that between 4 (10) and 78% (11) of those infected may be asymptomatic.

Another limitation is the bias created by the selection of employees as the target population when compared to the general population. The employees were regularly employed people with at least an elementary education who continued to work during the quarantine and who had differentiated access to information.

## Conclusions

The contamination of employees at our hospital reached high levels in April but was controlled in May. In-hospital surveillance may provide a proxy for understanding epidemic behavior in cities.

## Data Availability

The data is available upon request

## Data Availability

The data is available upon request

